# Machine learning analysis of Autism phenotype data supports a four-dimensional continuum with three overlapping subtypes

**DOI:** 10.64898/2026.08.27.26361561

**Authors:** Hugh Quigley, Bryan Gardiner, Liam McDaid, Cian O’Donnell

## Abstract

Autism Spectrum Disorder (ASD) is a heterogeneous neurodevelopmental condition defined by differences in social communication and restricted, repetitive behaviours. As diagnostic criteria have broadened, ASD is now recognised across a wider range of individuals, raising key questions about its structure: does ASD have discrete sub-types, or is it better conceptualised as a continuous, possibly multidimensional, condition?

We aim to explore whether a multidimensional continuum model more accurately captures the variability within ASD. We analysed a large SPARK phenotypic dataset of medical history and diagnostic surveys (background history, SCQ, RBS-R; n=36,710 individuals). We apply and compare two traditional statistical approaches, Factor Analysis and Gaussian Mixture Models, with a modern machine learning technique, the Variational Autoencoder (VAE).

VAEs reconstructed unseen test data with ∼4-fold better accuracy than Factor Analysis, and ∼8-fold better accuracy than Gaussian Mixture Models. We identified four stable latent factors across 100 independently trained VAEs. These four dimensions provide an individual behavioural profile that can be visualized using radar-plots, offering a compact way to compare profiles at the person level. Through further analysis, we found evidence for 3 overlapping clusters or subtypes of ASD identified within the 4D latent space. This work aims to inform new ways of modelling ASD using a VAE that will be able to discern between a continuum or a clustered output and that go beyond binary diagnosis, instead reflecting the complex range of trait profiles, with implications for personalised diagnosis and intervention.

**Lay Summary:** Autism varies considerably between individuals, making it difficult to fully describe using a single diagnostic category. Using machine learning on a large dataset of Autism survey data, we identified four continuous dimensions of autistic characteristics and three overlapping subtypes, highlighting that individuals can share characteristics across groups rather than fitting into distinct categories. These findings may help provide a more individualised understanding of autism and its diverse characteristics.

## 1. Introduction

Autism Spectrum Disorder (ASD) is a neurodevelopmental condition characterised by persistent deficits in social communication, the presence of restricted and repetitive behaviours, and a strong preference for routines and predictability (Hirota & King, 2023). Typically diagnosed in childhood, ASD is increasingly recognised across the lifespan due to evolving diagnostic criteria that now encompass a broader range of symptom presentations (Roy & Strate, 2023). While this shift has made diagnosis more inclusive, it has also introduced new challenges in how ASD is conceptualised and understood.

The rising number of diagnoses, particularly in individuals with less severe but still impactful traits (Maenner, 2023), raises important questions: Is ASD best represented as a discrete category or a continuous spectrum? Should it be viewed as a single-dimensional condition, or does it involve multiple distinct but overlapping domains of difficulty?

One concern with current diagnostic approaches is that they often fail to capture the specific traits that affect individuals uniquely. As a result, many people with ASD report that generic categorical labels do not accurately reflect their lived experiences (Charman et al., 2009; Giles, 2014). This lack of individual resolution can make it difficult for clinicians to allocate support effectively, as people with the same diagnostic label may require very different interventions, risking mismatched care that either under-supports high-needs individuals or overlooks strengths that could be developed. Instead, there is growing interest in conceptualising ASD as a multidimensional continuum. While this idea has gained support from the neurodiversity movement and some clinical perspectives (Gensler, 2012), there has been limited quantitative investigation into whether such a structure is supported by real-world data.

The current diagnostic framework for ASD can be characterized by the idea of a one-dimensional continuum (Figure 1 A) with a single axis of severity, which does not capture the true heterogeneity of ASD (Astle et al., 2022). Consequently, researchers have proposed alternative models, including discrete subtypes (Figure 1 B) (Grzadzinski et al., 2013) and a multidimensional continuum (Figure 1 C) (Lyall, 2023; Robinson et al., 2016).

**Figure 1:**
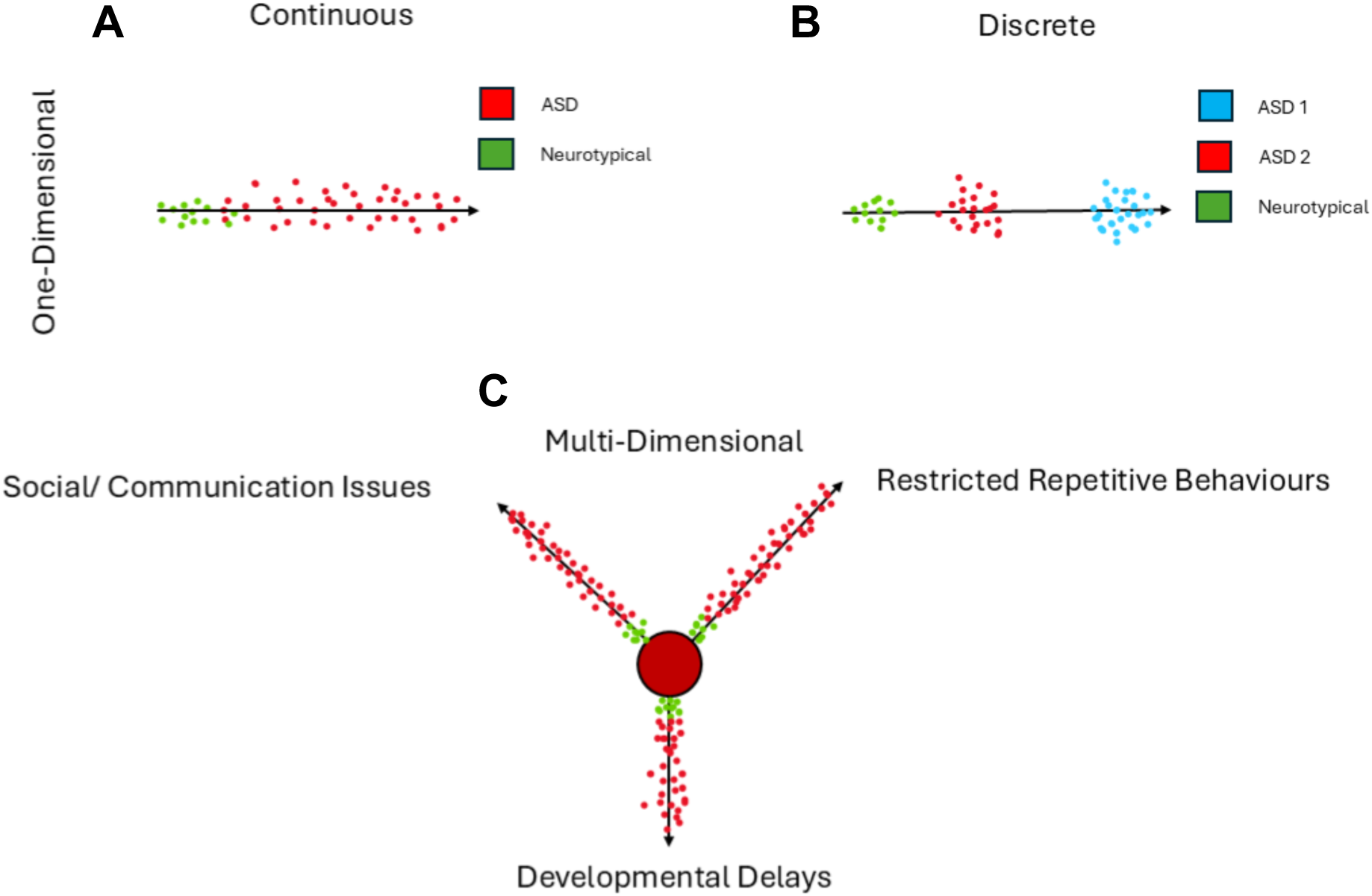
Dimensional vs categorical descriptions of autism. **Top left:** a possible one-dimensional continuum data distribution. **Top right:** a possible one-dimensional discrete clustered data distribution. **Bottom:** a possible multi-dimensional continuous distribution of the data based on the guidelines set for ASD diagnosis in the DSM-V.

The open question remains, what is the true symptom structure of autism? We address this by using modern data science techniques to examine whether ASD is better characterised as 1) a single– or multi-dimensional and 2) discrete-subtype or continuum construct. This approach is inspired by similar work in other neurocognitive conditions— for example, the model of schizotypy in schizophrenia, which captures trait variation along positive, negative, and disorganised dimensions (Kwapil & Barrantes-Vidal, 2015).

By exploring the underlying structure of autistic traits, this project seeks to inform the development of more accurate, flexible models of ASD that better reflect its complexity.

## 2. Methods

All plots and analysis were done in Python. The GMM and Factor Analysis models were implemented with the `sklearn` package, the VAEs were created with `keras` and `tensorflow` packages, and the `diptest` package helped with cluster analysis.

### 2.1 Dataset and Preprocessing

To assess the dimensionality and discreteness of ASD, we analysed phenotype data from the 2024 release of the SPARK dataset (Feliciano et al., 2018), which was collected over 31 research clinics across 26 different states in the US. In total the dataset contains information on 390,466 individuals, 162,238 of which have an ASD diagnosis, including both adults and children. Exact breakdowns of age and number of people can be seen in Table 1.

**Table 1:** Dataset information.

| Age group | Total number of people | Number of Males | Number of Females |
| --- | --- | --- | --- |
| 2-5 | 11,200 | 8545 | 2655 |
| 6-9 | 11,526 | 8959 | 2567 |
| 10-13 | 8514 | 6566 | 1948 |
| 14-17 | 5313 | 3925 | 1388 |
| 18-21 | 119 | 94 | 25 |
| 22-25 | 28 | 23 | 5 |
| Total Males: | 28,112 | Total Females: | 8588 |

We chose datasets that fit with the current diagnostic framework of the DSM-V: Social Communication Questionnaire-Lifetime (SCQ), Repetitive Behavior Scale – Revised (RBS-R), and the addition of the background history to try and get better view of ASD. This gave a base of 48,759 people to work with before further data cleaning.

During preprocessing, the merged dataset was examined for missingness across all variables. To preserve as much data as possible while maintaining completeness, all variables with more than 2,955 missing entries were removed. This threshold retained 94 variables and 36,710 individuals for analysis.

After variable selection, the remaining data was normalized using min–max scaling so that all values were transformed to a 0–1 range prior to model training.

### 2.2 Gaussian Mixture Model (GMM)

To determine the optimal number of clusters, Gaussian Mixture Models (GMM) were trained with 1 to 15 components. Performance was evaluated using log-likelihood that can be seen in Figure 3D. AIC and BIC scores calculated on held-out validation data were also calculated (Supplementary Figure 1). All the scores showed that best number of components for the GMM was 4 clusters. It is important to note though that GMMs may claim that there clusters, even in cases where there may not be distinct separations in the data (McLachlan & Rathnayake, 2014).

Log-likelihood measures how well the model explains the data, higher values imply better fit. Both AIC and BIC both reward the model based on fit while also penalising complexity. AIC approximates the model and is more about prediction accuracy, whereas BIC has a larger penalty on complexity and is more about trying to find the true model fit. For both a lower score implies better fit.

During training, the data was split into a 80% training set and 20% test set to assess out-of-sample fit accuracy. 30 random 80/20 splits were used to mitigate training variability, with the log-likelihood being tracked across all splits and averaged.

### 2.3 Factor Analysis

Factor Analysis was performed to identify latent dimensions underlying the ASD-related data. Log-likelihood was used to select the number of factors, tested from 1 to 20 (Figure 3E). Following the elbow method, while results get better with more factors, there is a slightly sharper incline from 9 to 10 factors. Therefore 10 factors were selected as a pragmatic choice rather than as the result of a strict optimisation criterion.

As with the GMM, we performed an 80/20 validation and training data split, repeated at random 30 times. Fit accuracy varied only minimally between runs, implying stable fit quality.

### 2.4 Variational Autoencoder (VAE)

To model complex, non-linear relationships within the ASD dataset, a Variational Autoencoder (VAE) was implemented to learn a low-dimensional latent representation of the input features. The VAE architecture consisted of a fully connected encoder, a four-dimensional latent space, and a mirrored decoder. Both the encoder and decoder had 3 layers. Dense layers with ReLU activation functions and batch normalisation were used throughout the network, with a sampling layer implementing the reparameterization trick to generate latent representations. The model was trained using the Adam optimiser for 50 epochs, with a beta value of 0.001 to balance the reconstruction loss and feature disentanglement. This objective encouraged the model to learn a structured latent representation while preserving reconstruction accuracy.

### 2.5 VAE factor consistency

When assessing VAE latent representations, we discovered differences in the factors between runs, presumably due to random weight initialisations and the stochastic training procedure. Given we aimed to find consistent factors for feature interpretability, we devised a procedure to discover consistent factors, at the cost of reduced reconstruction accuracy.

Figure 2A shows the consistency of factors for each latent dimension size over 10 runs per size; this heat map shows that the most consistent size over all dimensions is 4-dimensional run.

**Figure 2:**
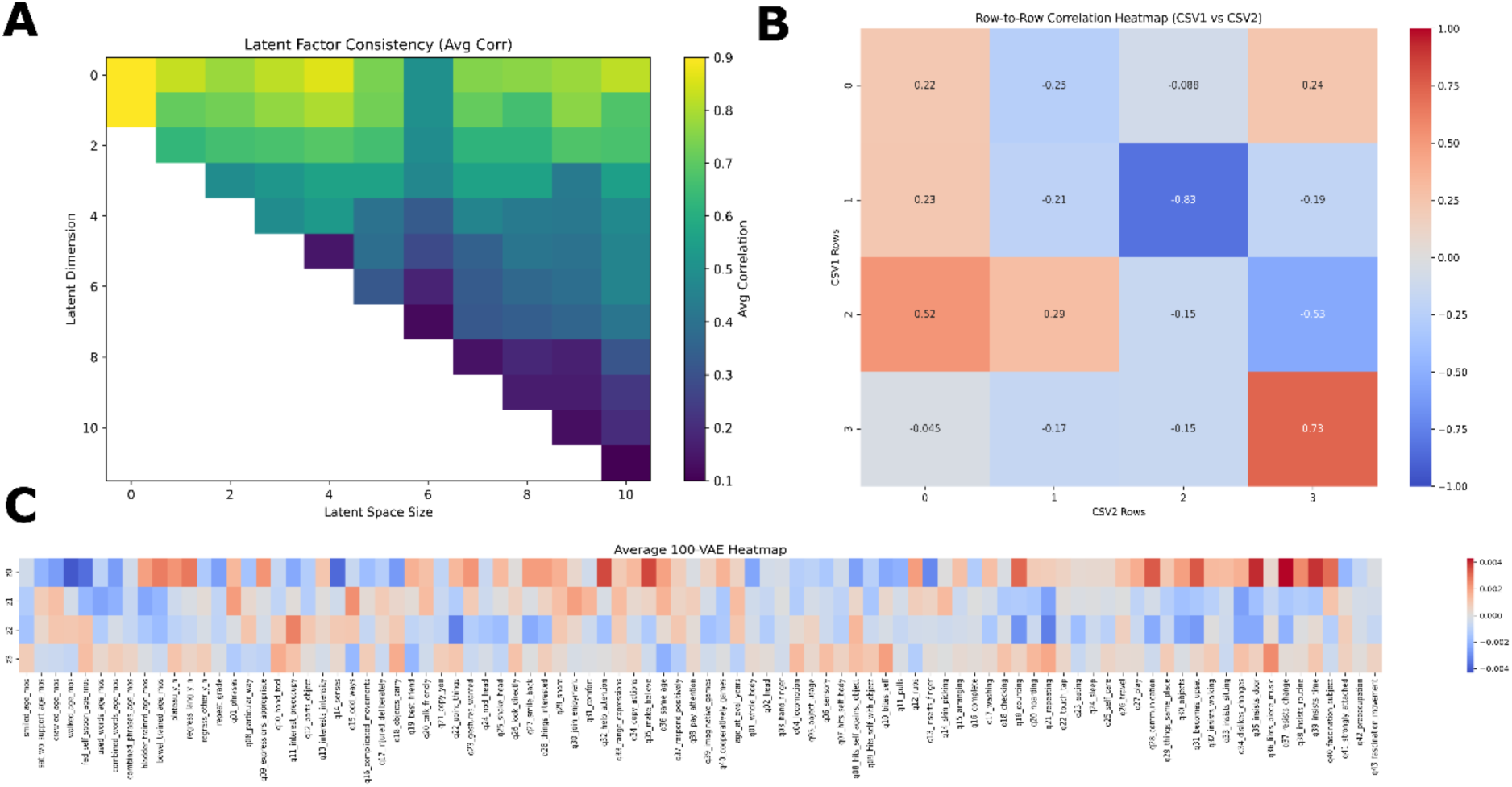
VAE dimension selection. **A:** heatmap showing factor consistency. **B:** Heatmap of correlation between 4-factor model representation and the top 4 consistent features from the 7-factor models. **C:** VAE feature heatmap showing 4 latent dimensions (rows) vs raw data features (columns).

**Figure 3:**
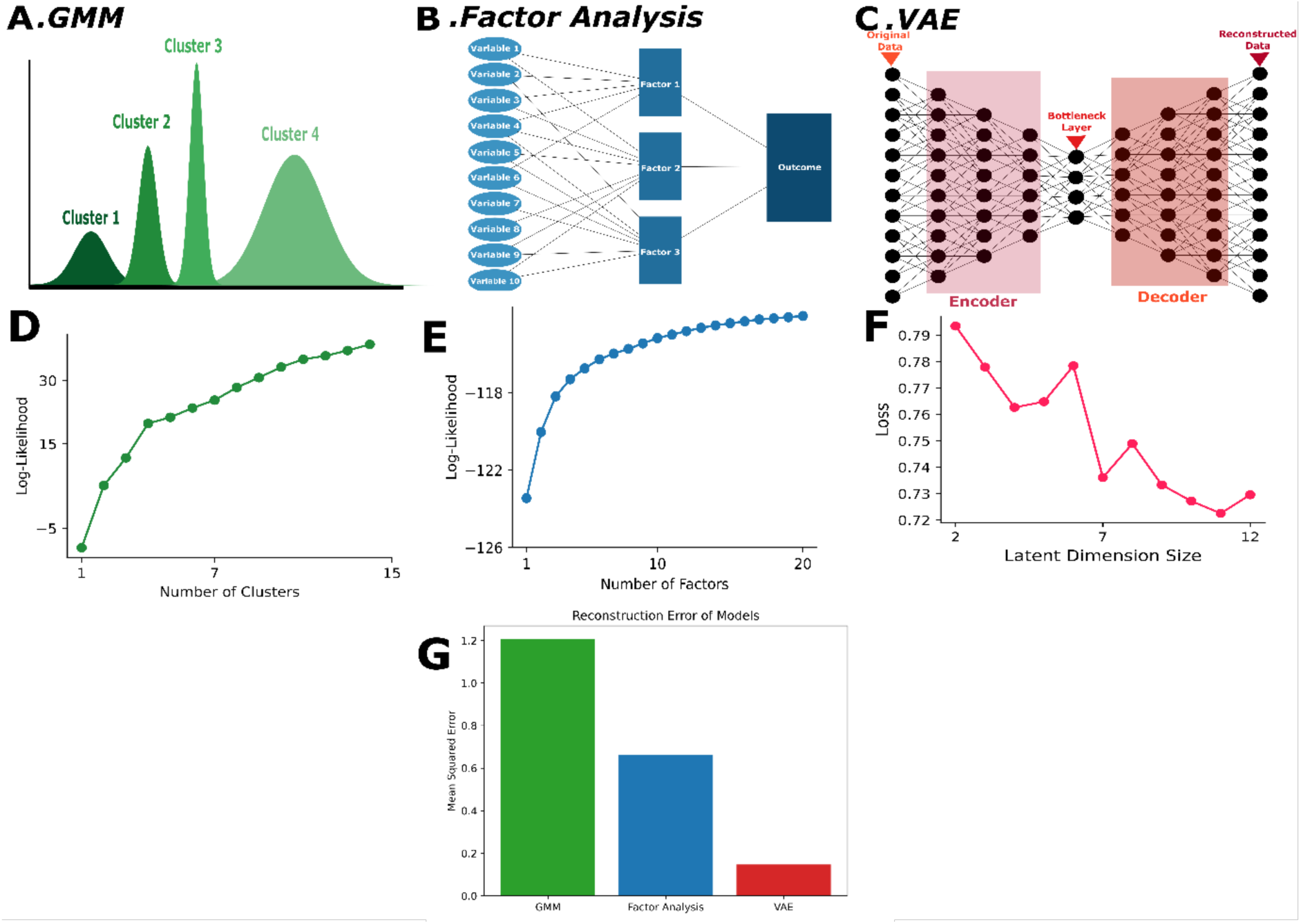
Model Architecture, Tuning, and Reconstruction Error across GMM, FA, and VAE. **A-C:** Schematic diagrams of each of the three statistical models: Gaussian Mixture models assume that the data arises from multiple separate clusters (A); Factor Analysis assumes that the high-dimensional data are explainable by a small number of linear factors (B); VAEs use deep neural networks to nonlinearly compress the data down into a small number of latent features (C). **D-F:** Goodness-of-fit measures on unseen test data for the three models (y-axes) vs number of latent components (x-axes). GMM plot (D) shows log-likelihood vs number of clusters; FA plot shows log-likelihood vs number of factors (E). Higher log-likelihood values imply better data fit. VAE plot shows loss vs latent dimension size (F). Lower loss values imply better data fit. **G:** Reconstruction accuracy mean-squared error (y-axis) of the three models on unseen test data.

With the new dimension size chosen there was still a need to validate whether the results from the 4 dimensions were also consistent with the 7 dimensions that showed best reconstruction accuracy. Figure 2B shows the top 4 most consistent factors in the 7-dimensional run and compares them to the 4-dimensional run. Their high similarity enough to validates the choice of 4 dimensions, in the trade-off between feature consistency and reconstruction accuracy. We therefore proceeded to use a 4-dimensional feature layer for the remainder of the study.

To assess the representations learned by the VAE, the encoder from each of the 100 trained models was used to project the unseen validation data into the four-dimensional latent space. Pearson correlation coefficients were then calculated between each latent dimension and each of the 94 input features across all individuals in the validation dataset. To account for variability in the ordering of latent dimensions between independently trained models, the latent dimensions from each run were aligned to those of the first training run using correlation-based matching. The aligned correlation matrices were then averaged across all 100 runs to produce a single correlation heatmap (Figure 2C). Notably there was some consistency in weightings between neighbouring raw data features in the heatmap – see adjacent reds and blues runs in each row. This reflects groups of survey questions that are thematically related, but the relationships were not confined to the three original surveys, showing a more complex pattern.

Radar plots were generated from the mean latent representation (z*_mean_*) of each individual, obtained by averaging the latent vectors across the 100 trained VAE models. To facilitate comparison between individuals, each latent dimension was normalised to the range using min-max normalisation:

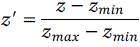

Where z is the average latent value for a given dimension, z*_min_* and z*_max_* are the minimum and maximum latent values across the individuals being compared. The resulting normalised latent values were displayed as radar plots using `Matplotlib`.

## 3. Results

To assess the dimensionality and distribution of ASD traits in population data, we applied three statistical models (Figure 3) to a large dataset of individuals with ASD phenotype measures (n=36,710, see Methods) (Feliciano et al., 2018; Fombonne et al., 2022), including a wide range of behavioural, developmental, and clinical scores. We compared two classical models as baselines: Gaussian Mixture Models (GMMs) and Factor Analysis (FA), with a modern nonlinear approach, the Variational Autoencoder (VAE) (Dai & Wipf, 2019). All three can be viewed as generative models of the population data but differing in their structural assumptions and representational capacity. Gaussian Mixture Models (Figure 3A) assume separable clusters and are useful for detecting subgroups, but struggle when boundaries are diffuse or nonlinear. Importantly, GMM fitting algorithms may claim that the optimal number of clusters are greater than one, even in cases where no separable clusters exist in the data (McLachlan & Rathnayake, 2014). Factor Analysis (Figure 3B), in contrast, assumes that the data can be explained by one large low-dimensional cluster. It reduces traits into continuous latent dimensions capturing shared feature covariance. However, FA assumes that these latent features are linearly related to the observation data, and so may miss complex or nonlinear structure in the data. Variational Autoencoders (Figure 3C) use deep neural networks to learn flexible nonlinear latent structure, so are capable of capturing more complex relationships in the data, but at the cost of higher computational and data demands. The large sample size of the SPARK dataset enables the use of VAEs for this problem.

Each model allows for a different approach to understanding dimensional structures of autistic traits. Our aim was to see how different models can capture the heterogeneity of ASD and if they provide meaningful clusters, latent traits or continuous dimensions that can allow for new diagnostic framework for ASD.

Each of the three models have a key parameter controlling the number of components that they assume underlie the observed data (Methods). We varied this parameter from 1 to 12 or more for each model looping through each hyperparameter most important for each model (GMM: number of clusters; FA: number of factors; VAE: latent dimension size), refitting the model separately each time, repeated over for 30 random subsets of the data. For all three models, the performance on unseen test data improved monotonically with increasing number of components, but with diminishing returns (Figure 3D-F). From a practical viewpoint for interpretability and clinical applications, smaller numbers of components are preferable. Based on these two competing demands of accuracy and interpretability, we selected an optimal number of components for each model: 4 for GMM, 10 for FA, and initially 7 for VAEs (Methods).

In order to compare the three models side-by-side we assessed their ability to reconstruct raw data scores from the compressed components representations. Figure 3G shows the mean-squared reconstruction errors. VAEs performed the best, FAs had approximately 4-fold higher error, and GMMs had approximately 8–fold higher error. Based on these results, we focus on interpreting and understanding the VAE latent components for the remainder of the paper.

Above we settled on 7 latent VAE components, however for this low-dimensional representation to be useful for clinical diagnosis, the features should be consistently discoverable in the data. When we assessed the consistency of these latent features across different random subsamples of the dataset and initialisations of the VAE optimisation, we found that only 4 of the 7 components had high consistency (Methods). As a result, we focus only on these 4 highly consistent components, with only a loss of ∼3% accuracy relative to the 7-component model.

### Interpreting the latent VAE factors

One potential use for this low-dimensional representation could be to support ASD diagnosis in clinics. For that application, these VAE components should be human-interpretable. However, interpreting the features of such reduced representations in VAE is challenging in general (Burgess et al., 2018; Higgins et al., 2017; Kim & Mnih, 2019). Our approach to interpretation was to find the raw data features that were most highly correlated with each latent component, as averaged over 100 independently trained and aligned VAE models. We thresholded the correlations at +/-0.0019 to identify the most consistently associated features for qualitative interpretation, then for each of the 4 components in turn, manually inspected the associated feature subset to develop a rational explanation for their grouping (Table 2).

**Table 2:** VAE latent components interpretations.

| Latent component | Umbrella term | Top 3 raw features | Description |
| --- | --- | --- | --- |
| 1 | Early Social Responsiveness vs. Repetitive Manipulation | ritualistic eating / mealtime<br>-0.002514<br>Need to repeat routine events<br>-0.003123<br>Was there ever a time when the individual lost other types of skills they previously had<br>0.002859 | This dimension contrasts early social–communicative responsiveness and basic postural milestones with repetitive, stereotyped, or atypical object-focused behaviour’s reflecting a split between emerging social engagement and restricted or repetitive behaviour patterns. |
| 2 | Sensory motor Independence | Bladder trained age | This dimension reflects functional motor and self-care development alongside heightened preoccupation |
|  | with Atypical Focus | months 0.002340<br>Does the individual ever injure themselves deliberately 0.002185<br>Spins, twirls, twiddles, slaps, throws objects 0.002103 | and self-injurious behaviour, suggesting an axis where increasing physical independence co-occurs with atypical interests or regulatory difficulties. |
| 3 | Behavioural Rigidity and Preoccupation vs. Social Imagination | Does the individual ever play imaginative games with another 0.002944<br>Preoccupation with parts of objects 0.002876<br>Likes the same CD, tape, record, or piece of music 0.002636 | This dimension is dominated by preoccupations, insistence on routines, sameness, and self-injury, paired with developmental markers, and strongly opposed to social reciprocity and imaginative play. It captures a classic rigidity/repetitive behaviour versus social engagement contrast. |
| 4 | Expressive Communication vs. Functional Development | Bowel trained age months - 0.002645<br>Has a normal range of expressions 0.002605<br>Spins, twirls, twiddles, slaps, throws objects - 0.002163 | This dimension contrasts expressive communicative signals with language, toileting, object-use, and social-numerical behaviours. It reflects a subtle dissociation between nonverbal expressive communication and broader functional or symbolic developmental skills. |

One potential usage of this low-dimensional representation could be to support a visualisation graphic for each individual’s autism profile, both to inform clinical diagnosis and provide a better understanding for the individual. In Figure 4 we show 4D radar plots for three example individuals from the dataset, where each component is mapped onto the four axes of a circle, with the distance from the origin along each axis determined by the normalised value of the latent component (Methods). A zero radius corresponds to behavioural factor not being present, and radius of one corresponds to severe behavioural factor (Methods). Each example individual shows a distinct radar plot shape. In general, these plots offer an at-a-glance characterisation of a given individual’s autism profile.

**Figure 4:**
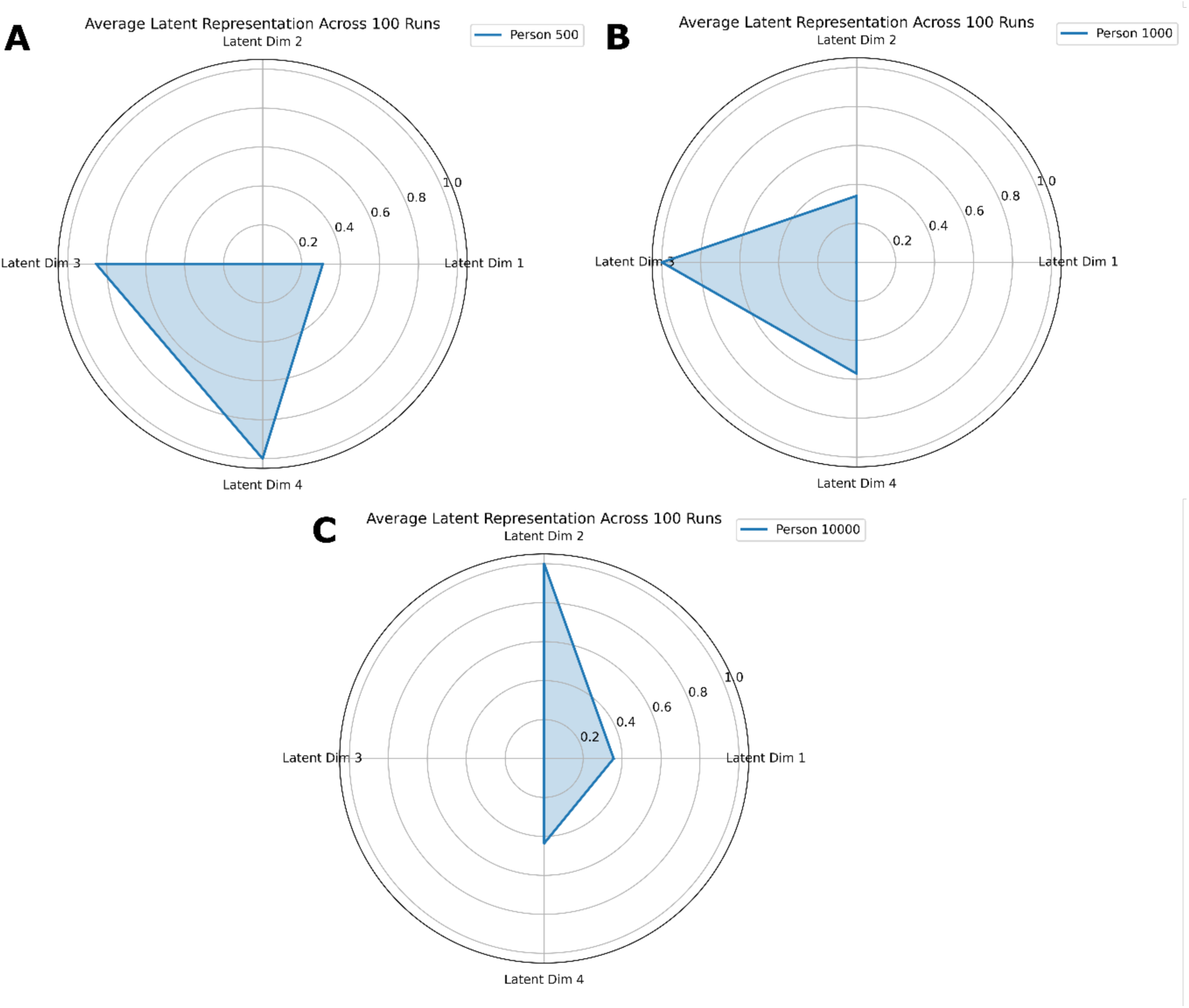
A-C: Example radar plots showing the 4D latent component profiles (Table 1) of three selected individuals from the autism phenotype dataset.

### Evidence for a multi-dimensional ASD continuum

To test if these data could distinguish between the discrete subtypes vs multidimensional continuum models of autism heterogeneity, we made histograms and pairwise scatter plots of the four latent component values for all 36,710 individuals (Figure 5). We reasoned that if discrete subtypes of autism phenotypes exist, they should be visible as distinct clusters or ‘bumps’ in these plots. Alternatively, there may just be one large continuum density.

**Figure 5:**
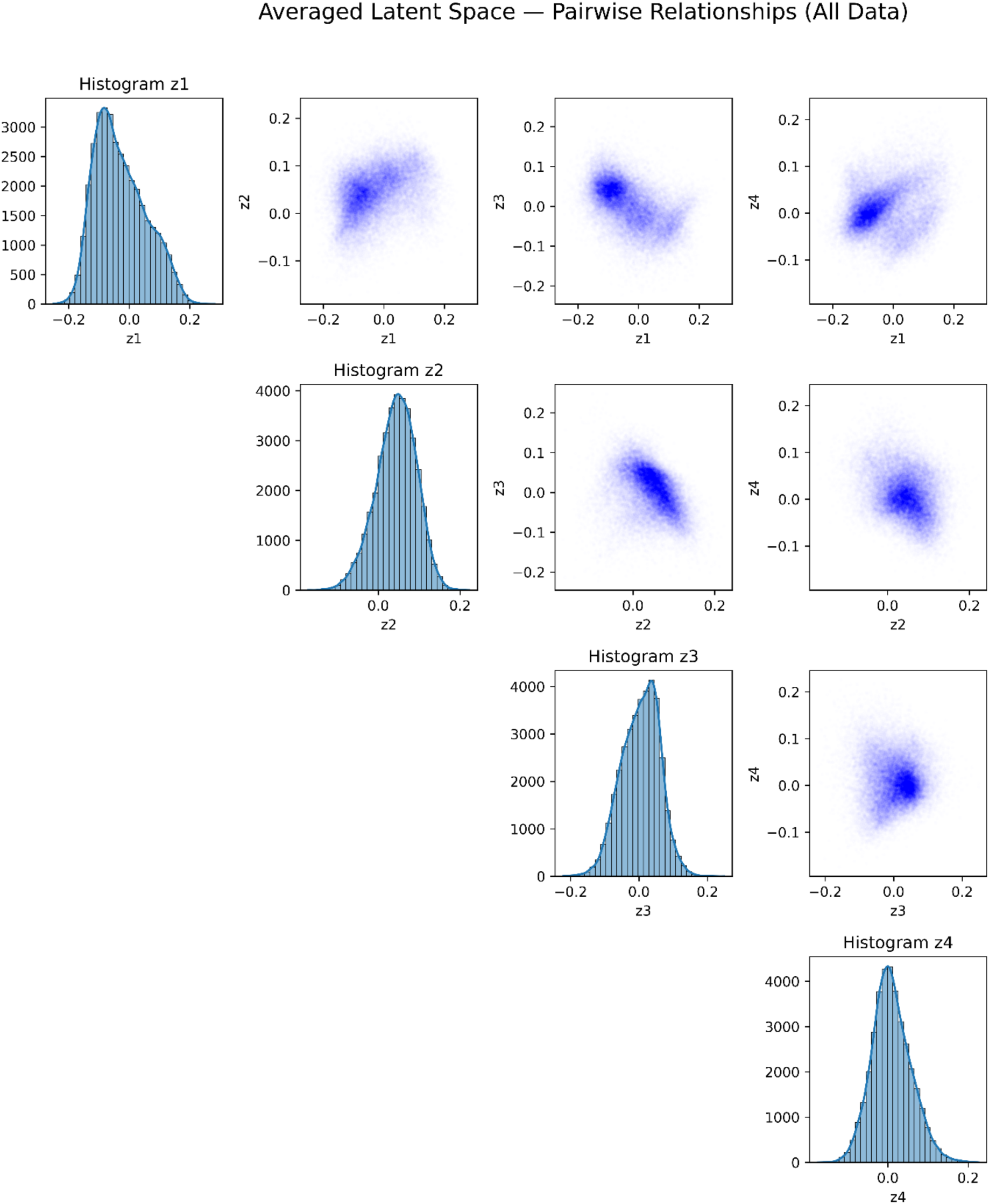
Scatter plots of all ∼36K individuals in our dataset, for each pair of latent VAE components, with histograms of each individual component on the diagonal.

Figure 5 is consistent with this multidimensional continuum model, as each scatter plot compares each dimension against one another with each dot representing a person within the data. Each dimension also has a histogram showing the distribution of the data within each of the dimensions, each histogram shows that this multidimensional model has no defined clusters within the data as each dimension has a smooth distribution of data.

To further test for clusters in the data, we used an automatic search procedure based on Hartigan’s dip test (Hartigan & Hartigan, 1985). This procedure takes many random 1D projections of the 4D data and statistically tests for multimodality in the histogram of the 1D projected data. If clusters exist in the data, these should be detected as a large fraction of low p-values. To validate this method, we first applied it to a well-studied 4D dataset of flower petal and sepal sizes (Fisher, 1936), which contains three species of flower that appear as distinct clusters. The line in Figure 6A shows the strongest dip projection (the most multimodal projection of the data) on two features from this 4D dataset, with projected values shown in Figure 6B. In this case, 91% of p-values < 0.05, indicating successful detection of multimodality in the data (Figure 6C).

**Figure 6:**
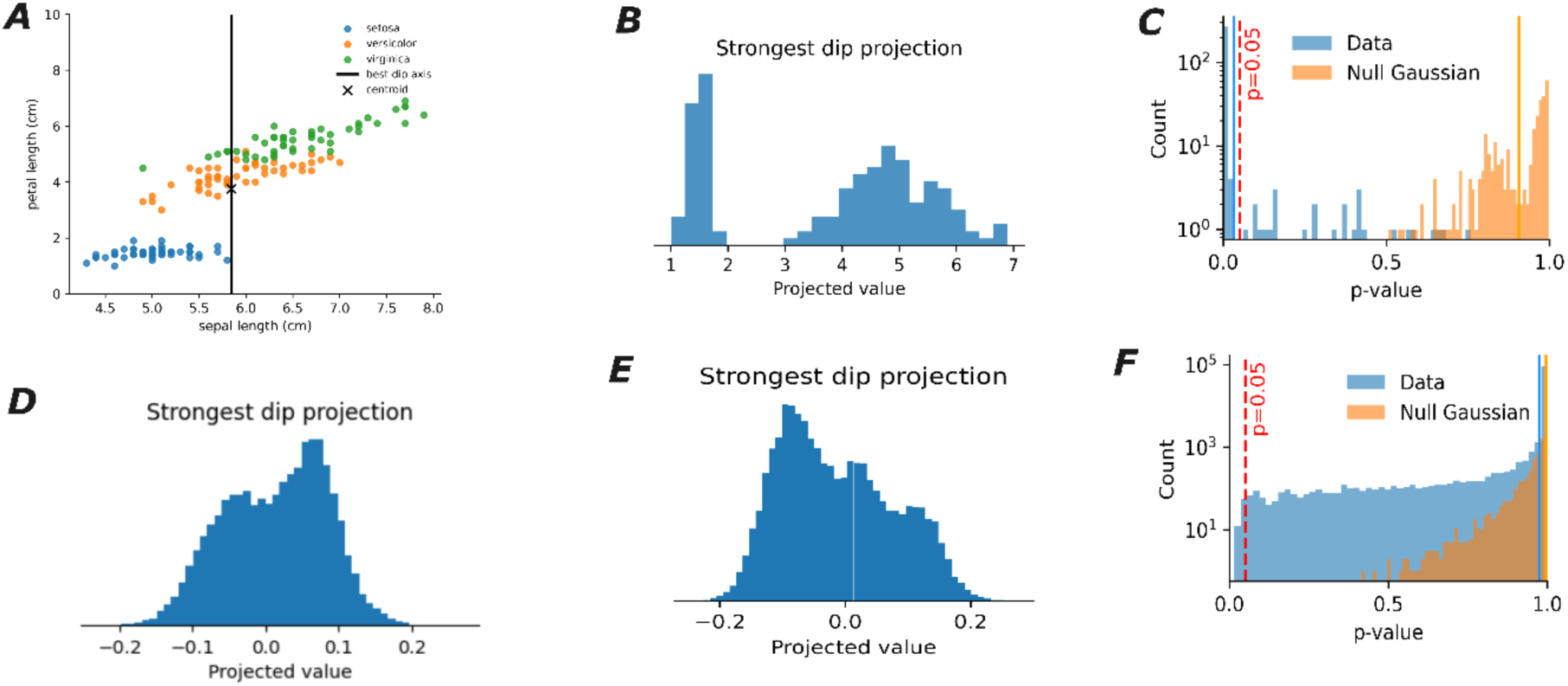
Dip test analysis. **A:** Dip test results for the iris dataset. **B:** Strongest dip projection from the Iris dataset. **C:** P-value histogram from the dip test search on 4D Iris data, showing strong evidence the multimodality. **D:** Strongest dip test projection from the 2-dimensional representations of the Autism data. **E:** Strongest dip projection of the 4-dimensional representation of the Autism data. **F:** P-value histogram from the 4-dimensional dip test search, showing evidence for multimodality in the data.

We then ran the same search procedure on each of the six pairs of 2D representations of our latent components (Figure 5, scatter plots). In this case, even the strongest dip projection (Figure 6D) did not show statistically significant multimodality (p=0.134). This implies that all 2-dimensional representations of the data were unimodal or clean continua.

Finally, we applied the same search procedure to our full 4D latent component data (Figure 6E, F). In this case, the strongest dip projection had 3 clear distinct peaks, implying multimodality (Figure 6E), with a large fraction random projections showing significant p-values <0.05 (Figure 6F). Although these clusters were not as distinct as in the Iris dataset, this nevertheless implies that there may be different clusters within the data that can be seem in the 4D representation and but not in 2D, showing at least three subtypes within the autistic individuals, that will be investigated further below.

### Developmental age correlates with Autism latent components

Age at diagnosis varied widely in our dataset (2-25 years). To test if age was correlated with the autism VAE latent components, we replotted the pairwise scatter plots colouring the markers by individual age at evaluation (Figure 7). Some dimensions show a clear gradient between younger and older individuals, especially Z1 and Z4. Linking this to our earlier interpretations of these components (Table 1), younger individuals tended to have a higher Z1 score, implying greater disparity between Early Social Responsiveness vs. Repetitive Manipulation. Younger individuals also tended to have higher Z4, implying a greater disparity between Expressive Communication vs. Functional Development than older individuals. Overall, these results support a developmental shift in autism profiles, and gives validity to the latent structure uncovered by the VAEs.

**Figure 7:**
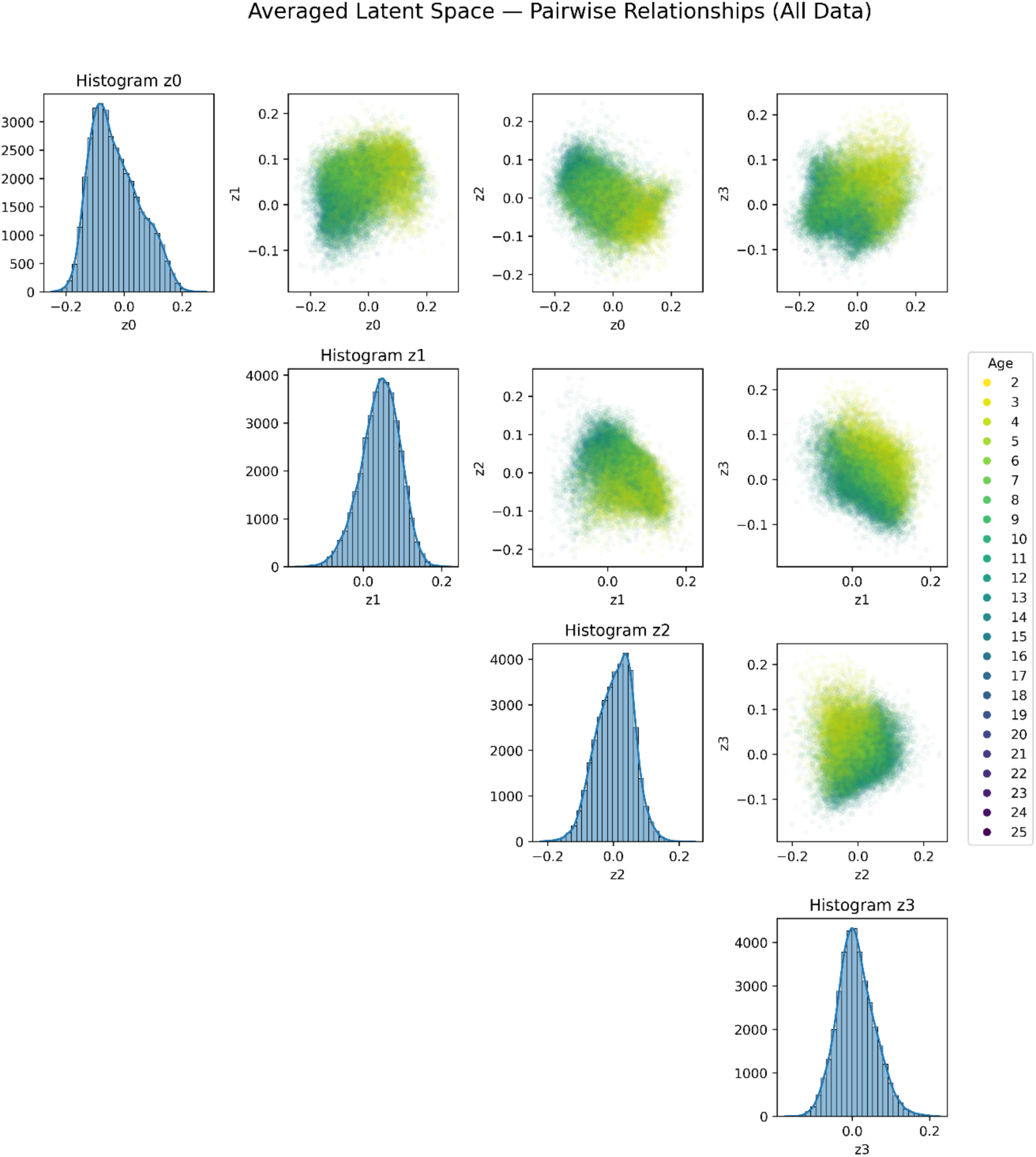
Similar to Figure 5, but with scatter plot markers coloured by the age (years) of each individual.

### Clusters in the 4 latent dimensions

Figure 6E and F showed some visual indication of 3 clusters within the 4 dimensions. We investigated this by running a 3-cluster GMM to detect the subgroups. To assess the quality of this clustering, we ran a logistic regression algorithm with cross-validation, and the clustered representation gave an 87.7% classification accuracy. To interpret these clusters we looked at the top 15 variables that distinguish the clusters (Figure 8B), allowing us to come to the following umbrella terms:

**Figure 8:**
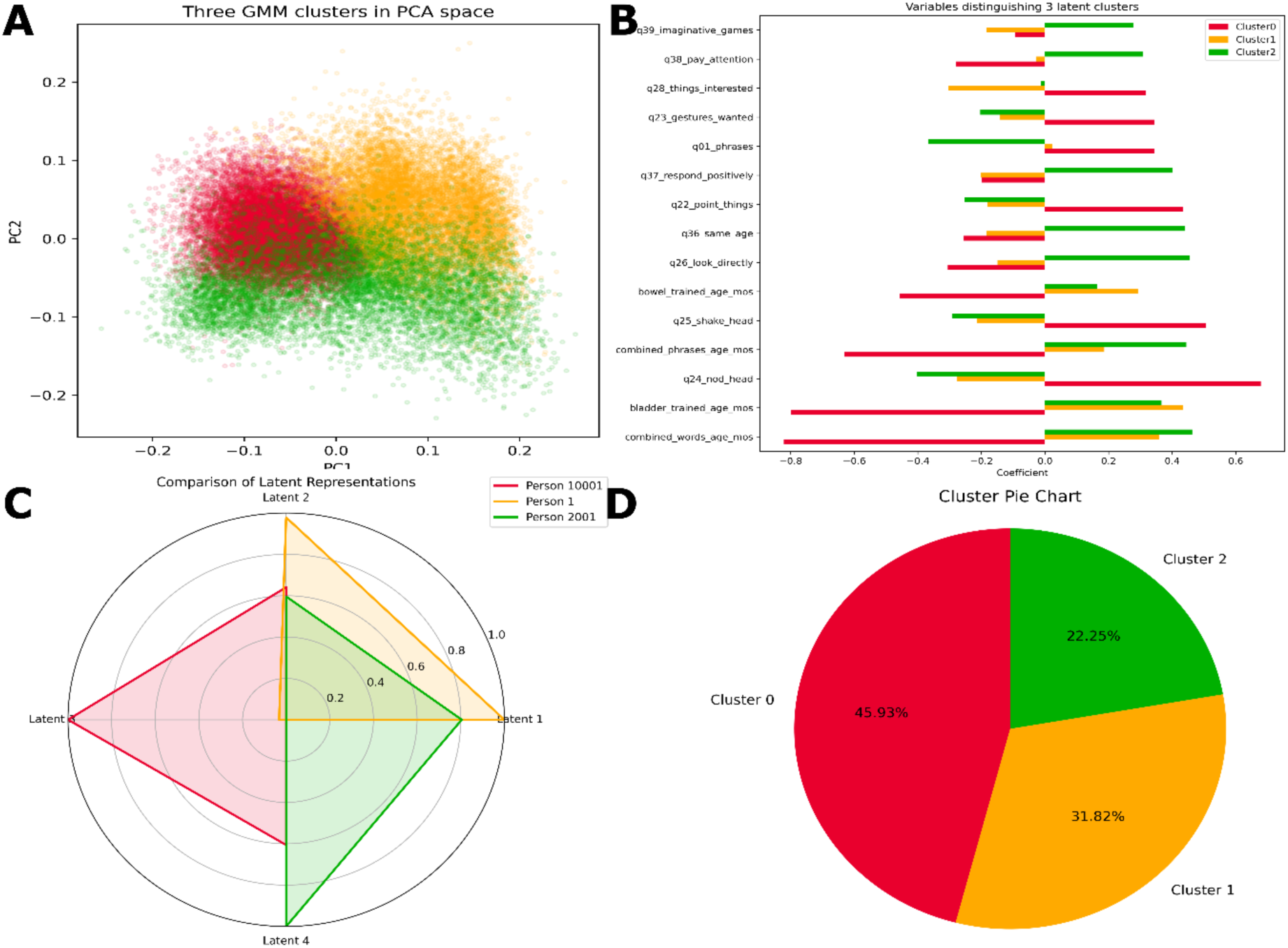
Clusters in latent space. **A:** PCA scatter plot of the clusters in the 4D space **B:** Top 15 variables for the three clusters, by GMM weights. **C:** Radar representations of the different clusters **D:** Pie chart showing the distribution of the cluster numbers.

Cluster 1 – Social Communication Impairment Profile – 16,857 people

Cluster 2 – Developmental and Language Delay Profile – 11,678 people

Cluster 3 – Social Interaction Impairment Profile – 8165 people

Figure 8C is a direct visual comparison of three example individuals that score highly in one given cluster probability score, but low in the other clusters. These radar representations showed the difference between the clusters and accounted for the unique factors in a way that is more discernible than the distinguishing variables (Figure 8B).

## 4. Discussion

Our results support a 4D multidimensional continuum model of Autism with 3 subtype clusters. This model offers a data-driven framework for profiling Autism symptoms, potentially allowing individuals and clinicians to better understand what parts of the vast Autism diagnostic criteria most directly apply to a given person.

Interpreting the latent factors discovered by the VAE inevitably required some subjective decisions (Methods), since the raw observational survey data was compressed into aggregate factors. It is possible that other researchers, clinicians, ASD individuals or family members could derive varying interpretations of the same latent factors. A survey of diagnostic tool’s target population could be done to establish a domain-appropriate consensus interpretation.

Here we used a modern machine learning method, the VAE, to compress the raw data into 4 latent dimensions. Although requiring large datasets to train, the extra model flexibility inherent in VAEs make them an appropriate technique for this type of problem, demonstrated by the improved reconstruction accuracy relative to traditional GMMs and Factor Analysis (Figure 3G). The hope is therefore that VAEs will discover more compact and subtle latent factors in the data, that capture the substantial heterogeneity of a condition like ASD. VAE has shown us that the dimensional output and clustered representations within the dimensions has a higher accuracy then the other models which allows us to assert that a multidimensional model of ASD is more accurate.

The VAE model carries some limitations. First, VAEs require larger datasets to constrain their parameters than simpler linear models like GMMs and FA. The recent availability of a suitably large Autism phenotype dataset [cite SPARK] made VAE use possible here, but it may prove challenging to apply VAEs to other Autism data types with smaller sample sizes, or toward smaller phenotype datasets for other neurodevelopmental or neuropsychiatric conditions. Second, VAE model design requires several researcher decisions about network architecture and training hyperparameters. While we systematically investigated the effect of various latent dimension sizes (Figure 2A), number of network layers, and learning rate parameters, an exhaustive search across all architecture and hyperparameter space was not possible. Third, interpreting the VAE latent space is known to be difficult since latent factors tend to be correlated across samples, and potentially unstable across training runs. While we addressed the latter problem (Methods), the former remains an active area of research (Li et al., 2025; Locatello et al., 2019; Zhou & Wei, 2020).

There were also some key caveats to our use of the dataset. First, while the dataset contained a mix of categorical and continuous survey data, our VAE assumed all raw data was continuous (Nazábal et al., 2020). Second, since were interested in within-autism heterogeneity and needed a large sample size, we restricted our analysis to individuals with an ASD diagnosis age 2-25 years old. Future analyses that include neurotypical individuals and individuals over 25 years old may uncover Autism signatures that we missed.

Here we analysed phenotype data only from individuals with a traditional Autism diagnosis, however multidimensional continuum models may also become appropriate for describing multi-disorder symptom structure. Indeed several neurodevelopmental and neuropsychiatric disorders have previously been proposed to span traditional diagnostic boundaries (Astle et al., 2022; Michelini et al., 2024; Owen & O’Donovan, 2017). The Hierarchical Taxonomy of Psychopathology (HiTOP), for example, aims to replace the existing categorical descriptions of psychiatric conditions in the DSM with a model that reflects the inherent heterogeneity and complex hierarchical structure of mental disorder symptom presentations (DeYoung et al., 2024).

Overall, our results support a multidimensional model of ASD that captures the heterogeneity of autism more effectively than traditional statistical approaches. The Variational AutoEncoder consistently identified four stable latent dimensions while achieving substantially better reconstruction accuracy than both Factor Analysis and Gaussian Mixture Models. Although the latent space was largely continuous, further analysis found evidence of three overlapping clusters within this continuum, suggesting that ASD may be better represented as a multidimensional continuum with regions of increased similarity, rather than completely discrete subtypes. From a methodological viewpoint, machine learning pipelines can provide a more detailed representation of autistic phenotypes and could support future intra-condition diagnostic frameworks by allowing individuals to be described through data-driven, quantitative trait profiles rather than a single categorical diagnosis.

## Data Availability

The phenotype data used in this study are not generically publicly available but may be requested for research purposes from SFARI Base.

https://base.sfari.org

## Acknowledgements

HQ was funded by a PhD studentship from the Northern Ireland Department for Economy. COD acknowledges funding from the UK Biotechnology and Biological Sciences Research Council (BB/W001845/1).

## Code and data availability

Python code for pre-processing the SPARK data, fitting the models, and plotting results is available at: https://github.com/hughquigley/Multidimensional_ASD_Code. SPARK Autism phenotype data is not publicly available but may be requested for research purposes from SFARI Base: https://base.sfari.org.

---

**Supplementary Figure 1:**
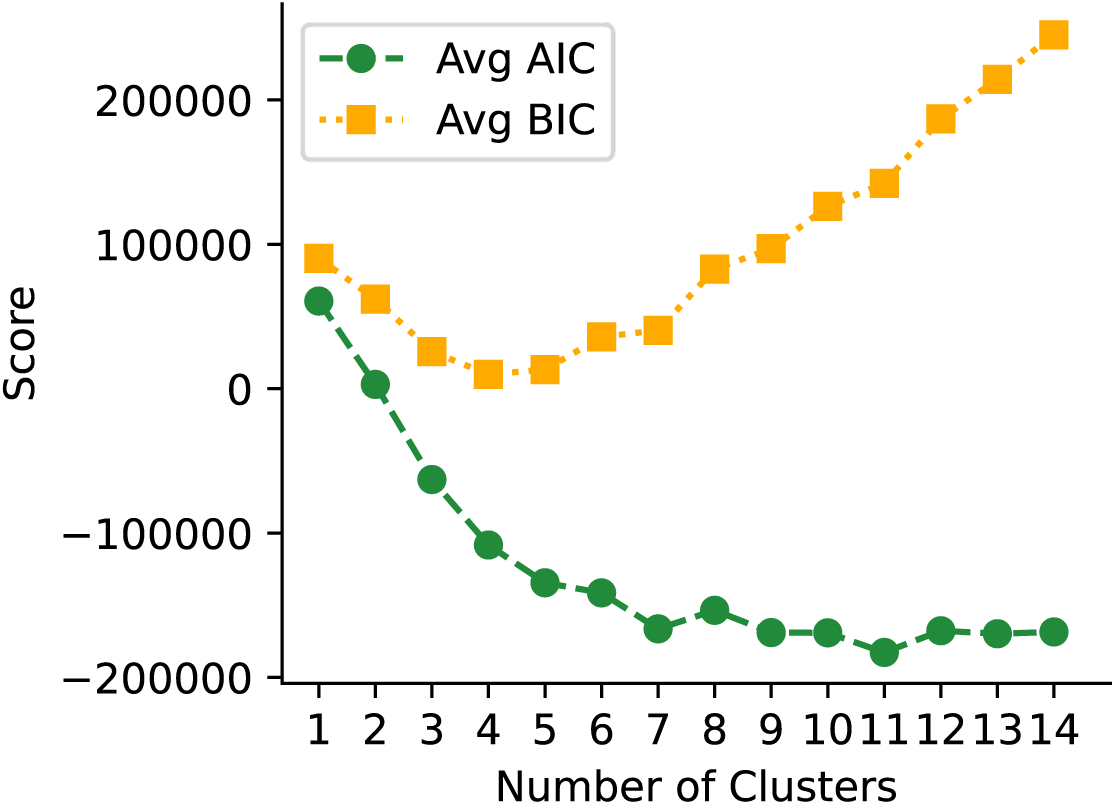
Average AIC and BIC curves for Gaussian Mixture model fit, as a function of the number of Gaussian clusters.

## Notes

### Competing Interest Statement

The authors have declared no competing interest.

### Author Declarations

Ethics committee of Ulster University Faculty of Computing, Engineering and the Built Environment gave ethical approval for this work.

